# Effect of control measure on the development of new COVID-19 cases through SIR model simulation

**DOI:** 10.1101/2020.10.27.20220590

**Authors:** Clara L., Fei Liu

**Affiliations:** East Brunswick High School; New Jersey Science Academy

## Abstract

In December 2019, the outbreak of corona virus disease, also known as COVID-19, was first reported in Wuhan, China [1]. Within only one month, the disease quickly spread to the United States through the transmission of respiratory droplets released when an infected individual sneezes or coughs [2]. Throughout the course of 9 months, the US reported over 8 million cases and 204,000 deaths, affecting the daily lives of American citizens [3]. As trends in corona virus cases are changing daily, it’s important to monitor these trends and observe the causes for the increase and decrease in new cases. The trends in new corona virus cases in the US as well as New Jersey are simulated using a modified Susceptible-Infected-Recovered (SIR) model. The new case graphs from the simulations reflect the new case trends in both the US and New Jersey and can be used to understand the mechanism behind the rates of corona virus infection as well as predict future corona virus trends. Comparisons between the results of the simulations and observed data show the effectiveness of control measures such as quarantine, physical distancing, and wearing masks. The extended time period of control measures taken in New Jersey led to a gradual decline in new cases reported daily while the US new cases showed a second wave of growth after control measures were implemented to a lesser extent.

## 1 Introduction

Corona virus is a novel disease that spreads faster than other contagious viruses like measles, and because most populations have little to no immunity to the disease, susceptibility to the virus is relatively high [4]. Subsequently, the virus can spread fairly rapidly, making the disease extremely dangerous; the number of people infected with corona virus can double or possibly even triple within a span of only a couple days as each infected individual is expected to pass the virus along to at least 2-3 others at early stages [5]. Subsequently, this infectious rate often results in an exponential graph of infected individuals, meaning once a small group of individuals becomes infected, the pool of infected people increases extremely rapidly. However, one other factor that significantly impacts the rate that individuals become infected are control measures like quarantine, social distancing, and wearing masks. When individuals wear masks, the risk of other individuals becoming exposed to contagious respiratory droplets that could contribute to the spread of corona virus is decreased. Because these control measures are implemented throughout the world, trends in corona virus cases are not always exponentially growing. In fact, the two samples that are studied in this paper, New Jersey and the United States, illustrate both rapid growth in new corona virus cases along with slow drops in new cases.

To study the mechanisms behind trends in corona virus cases, mathematical modelling is used to simulate reality. Research done by researchers Huppert and Katriel show that using mathematical modelling can be useful in providing valid predictions that correspond to reality [9]. Models are especially useful in filling gaps in the decision making process by utilizing already available data to predict future outcomes of pandemics.

To model fluctuations in corona virus cases, specifically, the SIR model is adopted. This model considers three main stages of infection: susceptible (S), infected (I), and recovered (R). Besides corona virus, SIR models have been used to promote interactions between decision makers and the modeling community through modeling diseases such as measles, norovirus, and influenza [6]. This specific mathematical model reflects human-to-human transmission as it models the flow of individuals across the three stages of infection: susceptible, infected, and recovered [7]. Generally, the performance of an SIR model is much better than an SEIR model, where E represents the exposed population, in terms of representing a set of confirmed cases, according to [8]. Work by this group of researchers show that although models like SEIR are more complex, predictions using these models are not necessarily more reliable than using simpler models like SIR. Moreover, SEIR models are more commonly used to model diseases with long incubation periods; as corona virus’s incubation period is relatively short, it is more reasonable to use an SIR model.

In order to utilize the SIR model and accurately depict the different stages of infection, it is important to note that the SIR model utilizes parameters and variables that can be altered. By altering these parameters, the results of simulations can be manipulated to reflect the results of reality. The parameters used in the equations are as follows: *N* = total population *α* = the transmission rate *β* = the average rate at which an infected individual can infect a susceptible one *λ* = the rate of recovery These parameters play an important role in producing graphs that accurately simulate the real-life confirmed cases of corona virus and are therefore essential to this research. However, it is also crucial to consider the limitations of using this specific SIR model. Ultimately, models are simplifications of reality, and therefore, in order to determine which simulations and predictions done by a model are reliable, it is important to do several runs and study a variety of outcomes [9]. Although the SIR model implemented simulates human transmission between different groups of infection, it does not take into consideration that individuals can become susceptible again after fully recovering from the infection. This is mainly due to the lack of data to conclude evidence of immunity to corona virus [12]. The SIR model used also assumes that control measures are implemented with the same effectiveness throughout each time interval. The purpose of this is to simplify the implementation of control measures when completing simulations; however, it is still possible to improve the implementation of this parameter when applying the model to a more realistic scenario [6]. To tackle these limitations and improve the accuracy of simulation results, two different regions are observed, New Jersey and the United States, and several methods of testing the reliability of a SIR model are used. Additionally, besides alterations to the already existing parameters, new parameters are implemented into the SIR model; this provides more flexibility in producing accurate results, one of which includes accounting for individuals becoming susceptible to the disease even after fully recovering. These adjustments will later be explored through detailed explanations of SIR model equations as well as in discussions regarding simulation results.

## 2 Extending the SIR model

We start from the traditional SIR model [6] [10] described by the following set of first order linear ordinary differential equations. These equations are extended to suit our modelling needs later on.

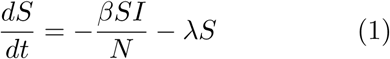

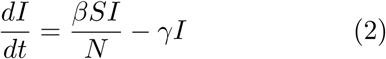

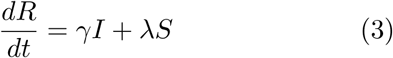

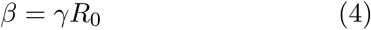

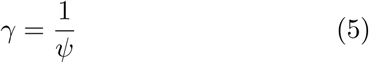

where *N* is the total population. Two important parameters that affect the transition between compartments are *γ* and *β*, where *γ* is the reciprocal of *ψ*, the infectious period (see equation 5). This is the period of time when the infected population can spread the disease to others, and the time is measured in days. For example, measles have an infectious period of approximately 8 days; this means between 4 days before and after rash onset, individuals can spread the disease to those around them. To explain the parameters in more detail, *γ* affects how the population transitions from being infected to recovered (see equations (2) and (3)). Next, *β*, also referred to as the “force of infection”, describes how quickly a disease can be transmitted through a population; ultimately, it is the product of *γ* and the reproductive number (*R*_0_), and affects how the population transitions between the susceptible and infectious groups (see equations (1) and (2)). In many cases, an infection will lead to a number of secondary infections, which is represented by the variable *R*_0_ (see equation (4)) [6].

As noted earlier, control measures play a significant role in the transition of individuals between the three SIR populations. This parameter, *λ* or control measure effectiveness, describes the fraction of individuals that are removed from the susceptible population. For example, when *λ* equals 0, this implies that control measures such as quarantine have no effect, and therefore, no individuals transition out of the susceptible group. On the other hand, if *λ* equals 1, this means that control measures are 100% effective; this would subsequently remove all susceptible individuals from the group [6].

This set of equations models the transition from *S* to 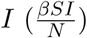, transition from *S* to *R* (*λS*), and transition from *I* to *R* (*γI*).

To make these equations easier to use, the functions are dimentionalized using total population *N* e.g. *s* = *S/N, i* = *I/N*

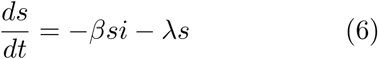

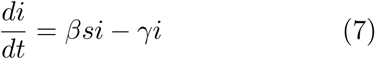

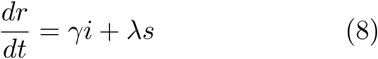

At the end of the model simulation, one can multiply *s, i, r* by *N* to recover the more realistic integer number of population in each pool.

To introduce the mortality effect, we add a new term that removes some of the infected to mortality pool by a proportionality constant *α*.

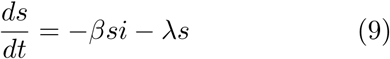

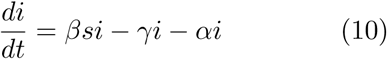

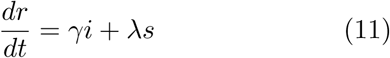

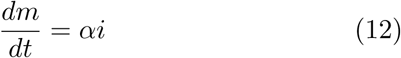

*α* is the mortality rate of the virus, and the current estimation is between 5-10% based on data reported from [11].

In order to model multiple infections, where recovered individuals can still become susceptible again, another term *ηR* is added to the *s* pool

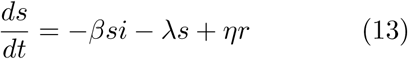

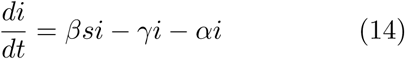

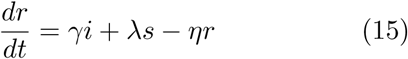

Note that the effect of multiple infections is much weaker, which is reflected as *η << λ. η* and *λ* carry different physical meanings but the numerical simulation simply adds the two effects together, and therefore, a single parameter can be used to model both effects *λ*^*1*^*s* = *λs − ηr*.

## 3 Discussion on the effect of parameters in the SIR model

First we examine the effects of the parameters on the SIR equations 13-15.

In the control run simulation (Figure 1), we used the following values for parameters. *β* is 0.08, *γ* is 1/14, *λ* is 0, and *α* is 0.

**Figure 1:**
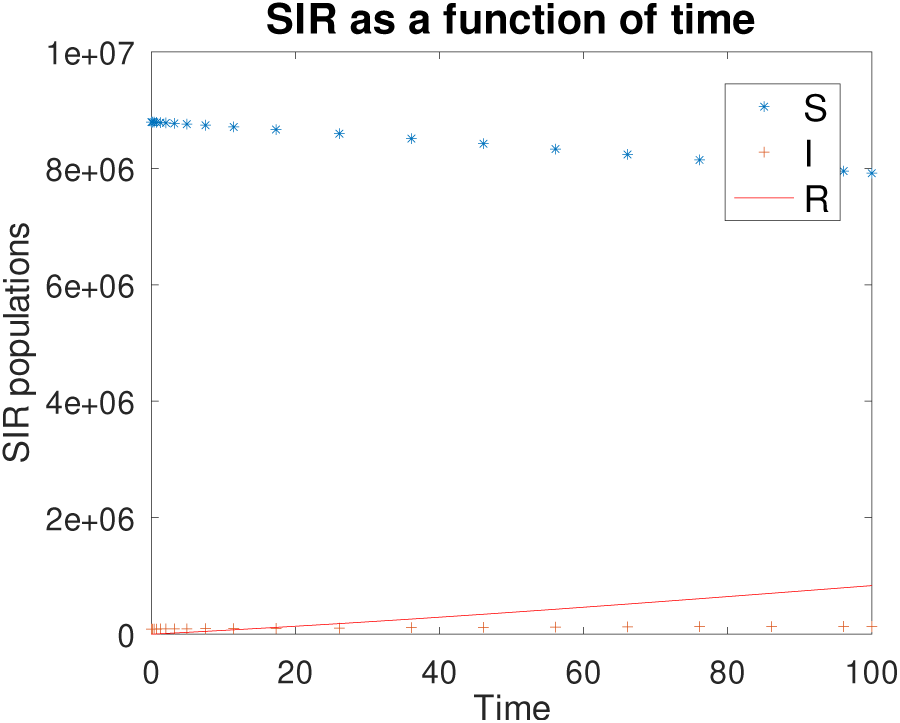
This is the control run result using the parameters specified above. The horizontal axis is measured in days and the vertical axis represents the total populations.

If we reduce beta from 0.08 to 0.04, we get the following results (Figure 2).

**Figure 2:**
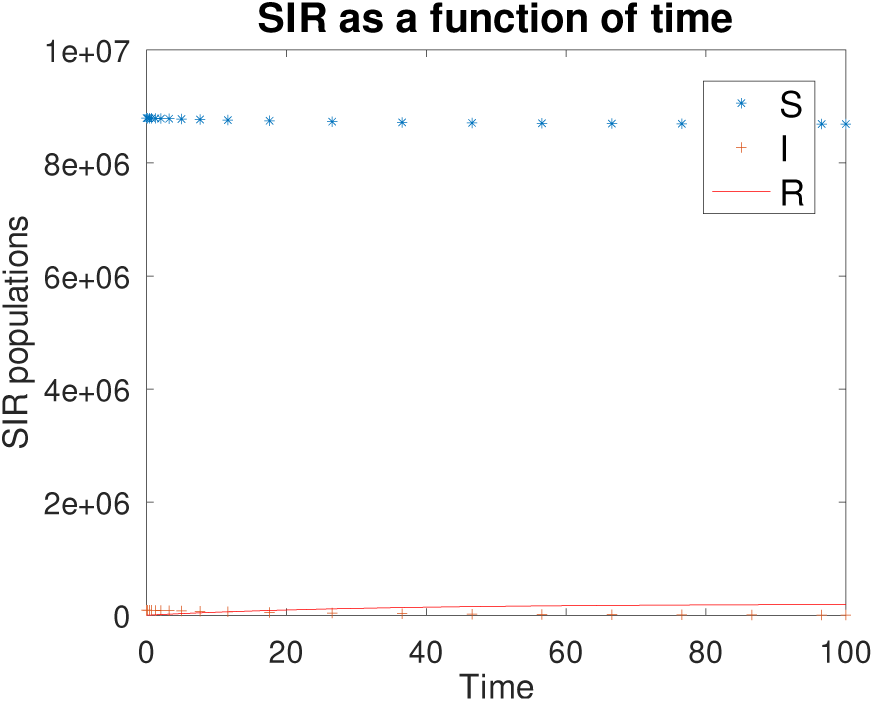
SIR when *β* is reduced from 0.08 to 0.04.

In this case, the rate at which susceptible people become infected decreases significantly. The curve is almost flat and is ultimately due to how quickly *β* affects the rate that people become infected.

If we reduce *γ* from 1/14 to 1/28, we get the following results (Figure 3).

**Figure 3:**
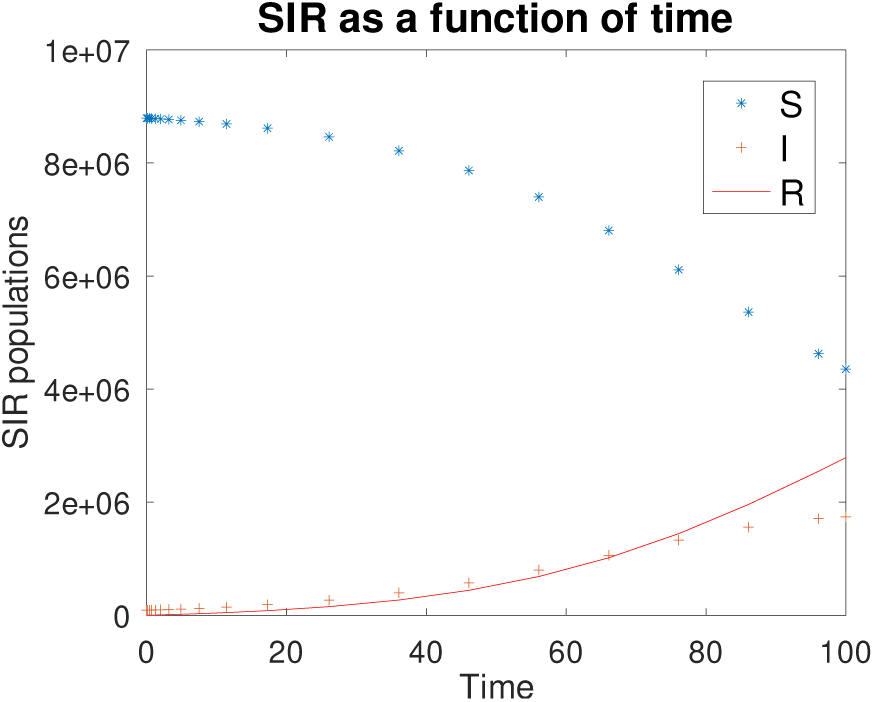
SIR when *γ* is reduced from 1/14 to 1/28.

When *γ* decreases,the rate at which infected recovering becomes smaller. This causes the number of infected people to become higher compared to the control run. This subsequently causes more susceptible people to become infected due to the larger number of infected people. Therefore, the rate at which the susceptible people that become infected increases.

If we increase *λ* from 0 to 0.005 (0.5% of susceptible people are transferred to recover per day), we get the following results (Figure 4).

**Figure 4:**
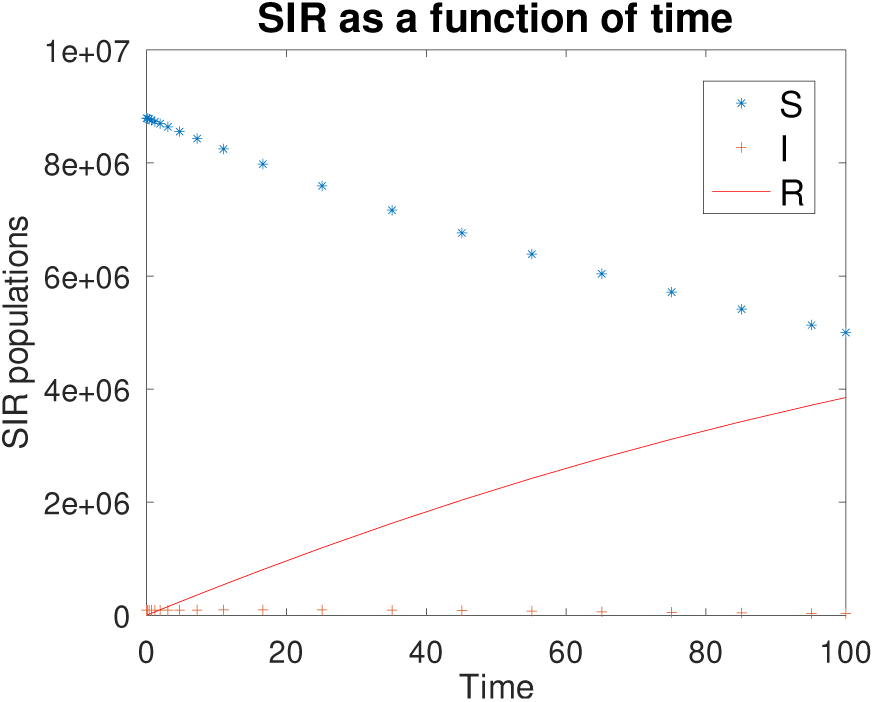
SIR when *λ* is increased from 0 to 0.005.

When *λ* increases, this represents the implementation of control measures on the susceptible population, whether through quarantining individuals or establishing social distancing regulations. With these implementations, the susceptible population is moved to the recovery pool, where they can no longer become infected. In the figure, we can also see that the infected pool is slightly decreasing because less people are susceptible to the disease, directly impacting and lowering the number of infected people.

If we increase *α* from 0 to 0.05, we get the following results (Figure 5).

**Figure 5:**
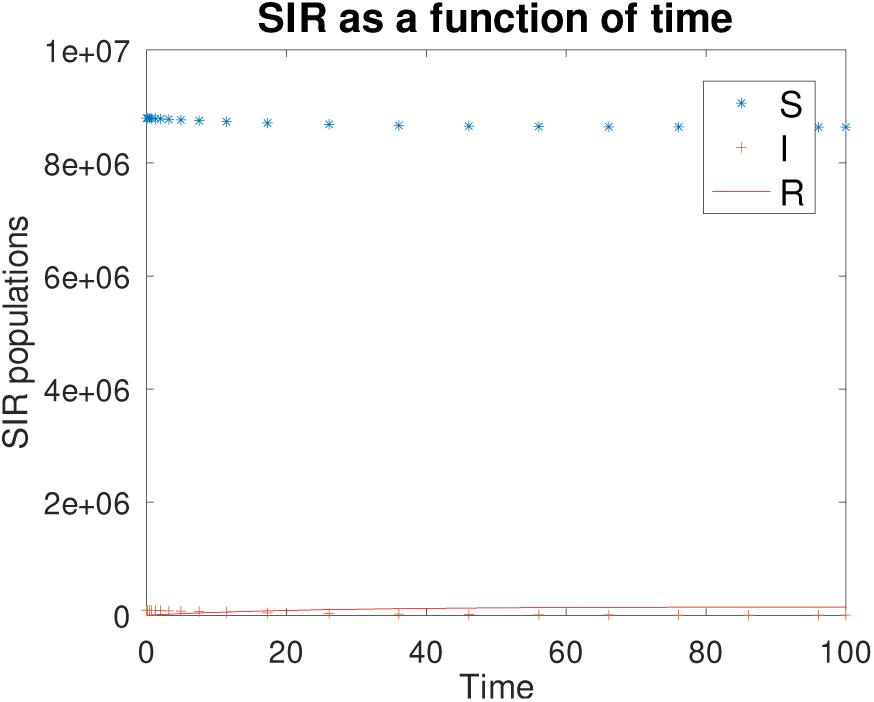
SIR when *α* is increased from 0 to 0.05.

Once *α* is increased, the rate of mortality caused by the disease increases. Consequently, the size of the infected pool becomes smaller while the size of the susceptible pool remains relatively the same because the deceased cannot infect them. The recovering pool also remains relatively the same because the infected people are dying instead of recovering.

The continuous dependence of the infected population on the various parameters are shown below: 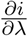 (Figure 6), 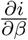 (Figure 7), 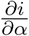 (Figure 8), 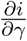 (Figure 9)

**Figure 6:**
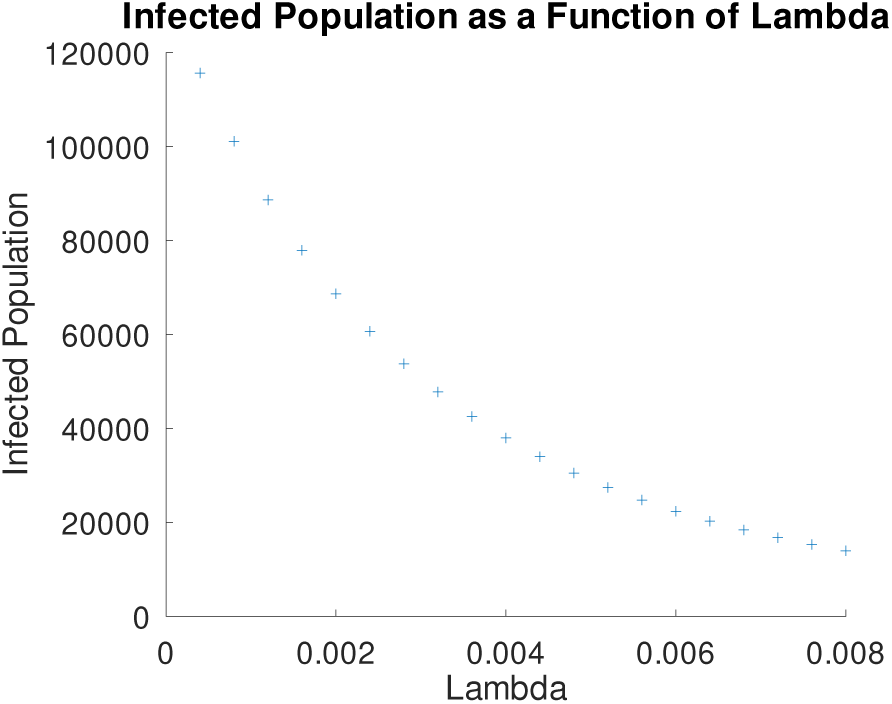
The infected (I) pool of people as *λ* increases.

**Figure 7:**
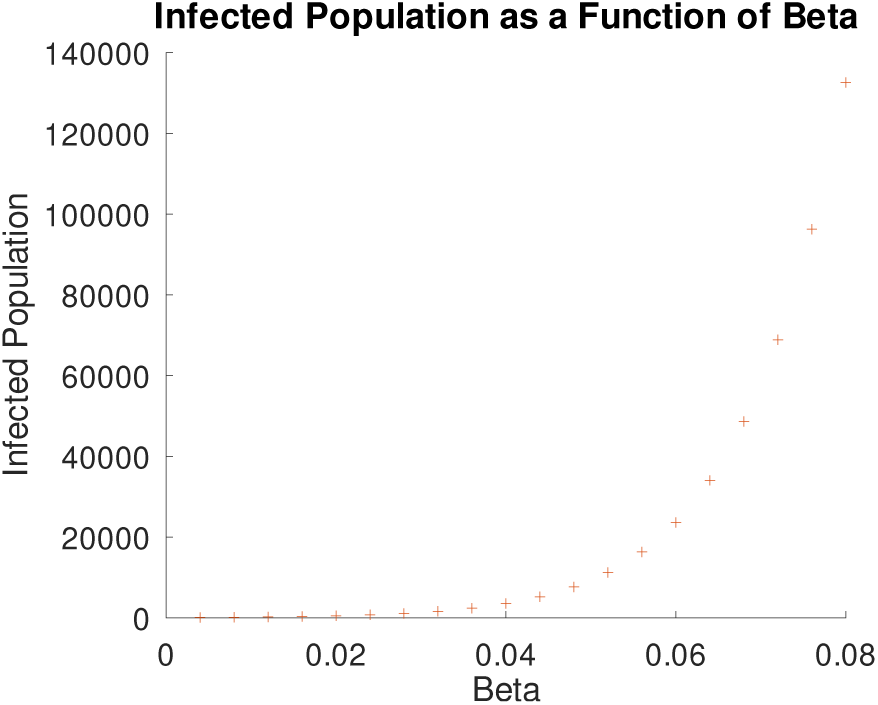
The infected (I) pool of people as *β* increases.

**Figure 8:**
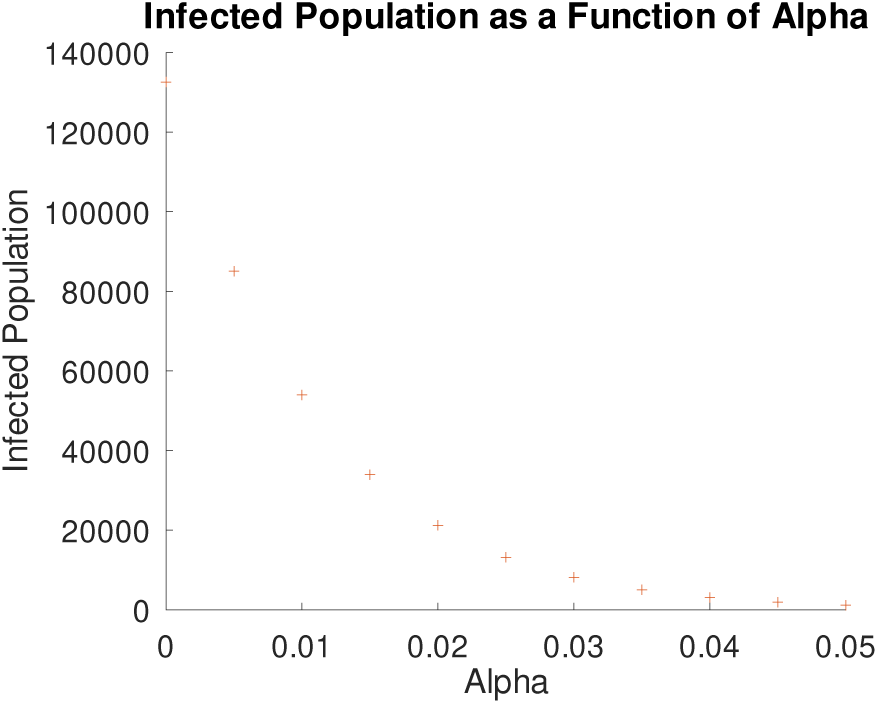
The infected (*I*) pool of people as *α* increases.

**Figure 9:**
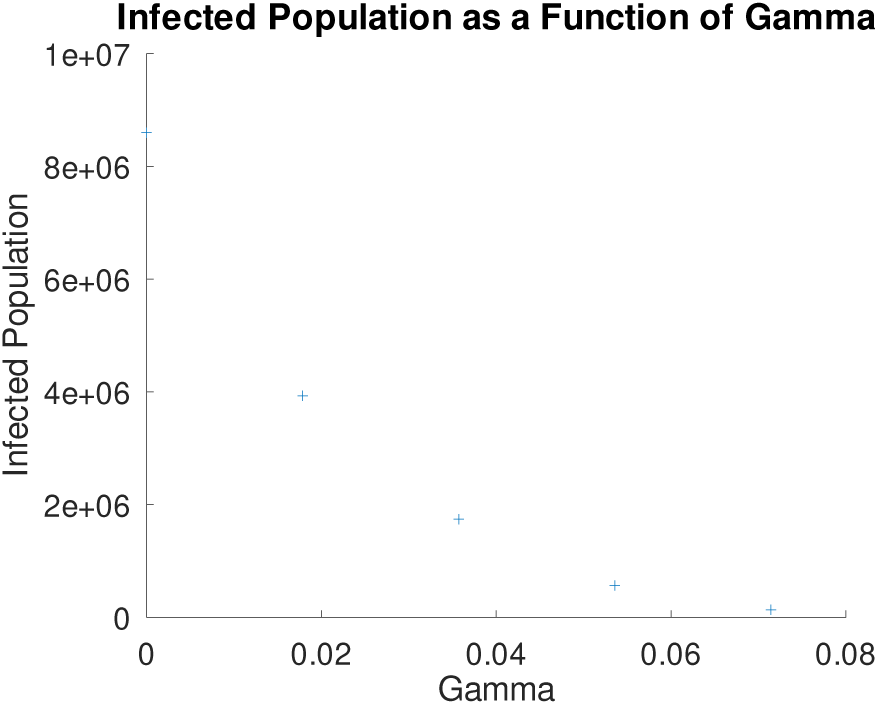
The infected (I) pool of people as *γ* increases.

*λ* represents the control measure effectiveness. Therefore, as this effectiveness increases, the population of infected individuals decreases. Control measures like quarantine assist individuals in faster recovery, thus resulting in a smaller population of infected individuals. However, the figure illustrates that while an increase in *λ* results in a rapid decrease in the infected pool of individuals from *λ* values of about 0-0.003, as *λ* increases more, the rate that the infected population decreases begins to slow down. This suggests that control measures have a limited effect on population size.

As *β*, the rate that people get infected, increases, so does *i*, the population of infected individuals. The figure above illustrates that values of *β* up to about 0.04 result in slow increases in the population infected people. Once past approximately 0.04, the slope of the infected population increases significantly, demonstrating an increased rate of infection. This implies that *β* has increased effects as the values get increase.

As *α*, the effect of death increases, *i* decreases as well because individuals from this population are dying. Moreover, as more people are dying, there is a smaller likelihood that individuals will get infected, causing *i* to decrease as well. The figure illustrates a rapid decrease in the number of infected people when *α* has lower values. As *α* begins to increase, the infected population decreases at a slower rate.

As the infectious period decreases, *γ* increases, so as *γ* increases, less individuals are likely to become infected. As a result, *i* decreases as *γ* increases; more specifically, the rate of decrease in the infected population is much higher at small values of *γ*, while at larger *γ* values, *i* decreases less rapidly.

## 4 Model results vs. Measurements

With the understanding of the dependence that *i* has on the various parameters, we examine how we could set up simulations to match what’s measured in New Jersey and in the entire US.

First, we graph (Figure 10) using the measured data from Johns Hopkins University (JHU) [11]. This continuous stream of data reports daily accumulated cases around the globe; the data is downloaded and processed in a spreadsheet. Note that because the JHU data set reports accumulated cases daily, one way to use the data is to calculate new cases by taking the difference between the accumulated cases every day.

**Figure 10:**
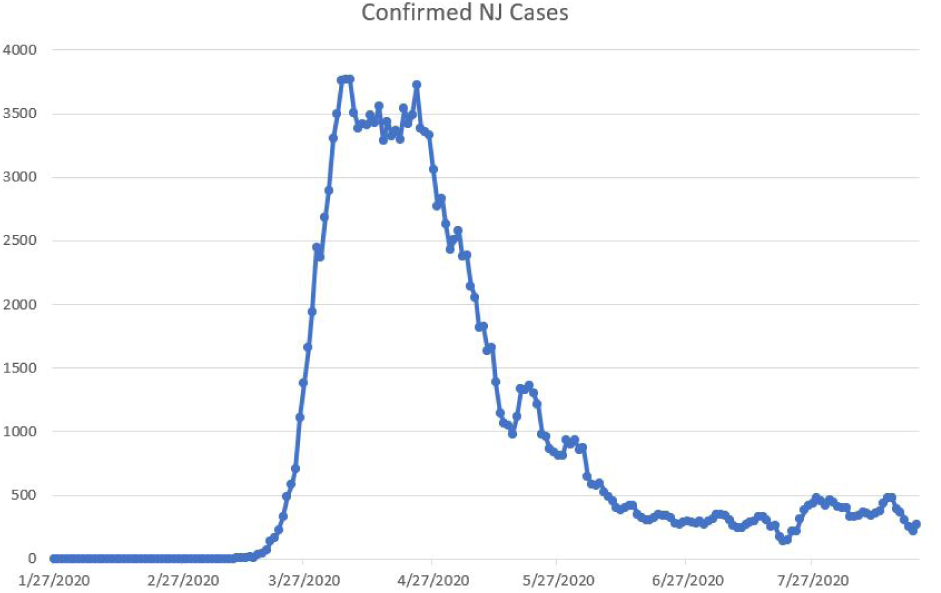
Graph of confirmed coronavirus cases in New Jersey from 1/27/20 to 8/21/20.

It can be seen that after an initial ramp up in the first 50 days or so, the reported new cases continue to decline steadily until it reaches a steady rate after about 100 days.

To simulate this steady decline more realistically, we introduce a time dependent control measure in the equation For New Jersey. A good candidate is a step function for which the sigmoid function is used.

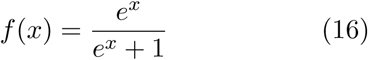

The sigmoid function has a nice property: it ramps up quickly near *x* = 0 and plateaus at *f* (*x*) =1. We can shift how this function influences the control measure by introducing a time delay and the slope at which the control measure kicks in by introducing scaling parameter *a* (Figure 11)

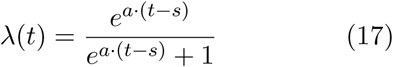

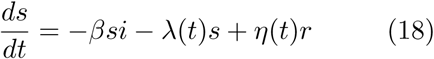

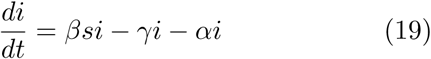

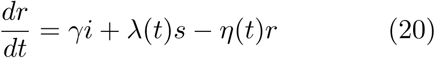

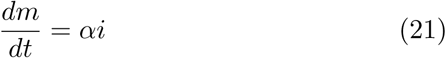

where *η*(*t*) is the rate at which recovered people can become susceptible again. A scaled exponential decay function is used to simulate 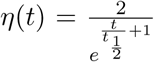 where *t*_1/2_ represents how quickly *η* decays over time.

**Figure 11:**
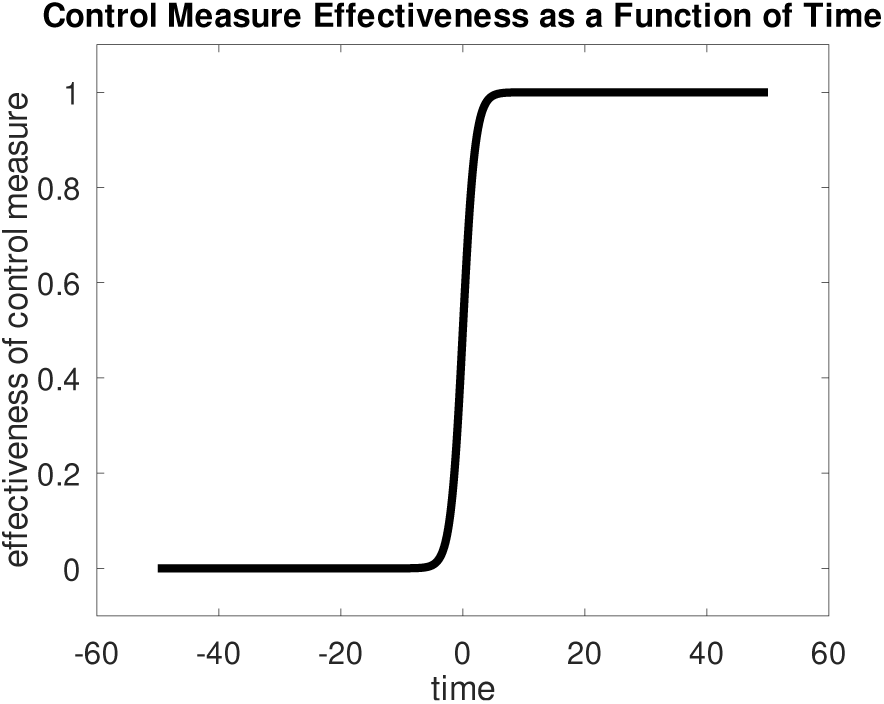
Sigmoid function (Equation 17) used to model the effect of control measure in NJ since the new cases in NJ plateaus after one month and decline sharply after two months. The sigmoid function behaves as a step function that simulates persistent control measure effect after it is introduced.

**Figure 12:**
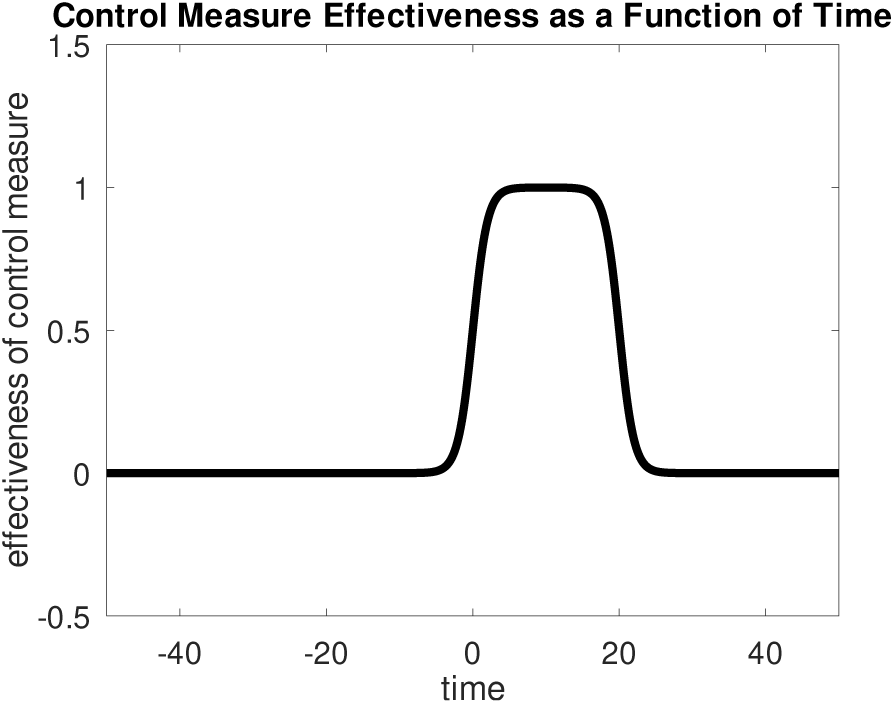
Rectangular function (Equation 23) used to model the effectiveness of control measures in the US. The parameters and their corresponding values used are *s* = 0 and *w* = 20 days. In this example plot, the control measure only takes effect during the first 20 days using this rectangular function.

Equation 18-Equation 21 represent the final set of equations in the SIR simulation in this work.

For the entire US, the new case pattern with a second wave of growth suggests it is better modelled by a rectangular function for the control measure by combining two sigmoid functions together

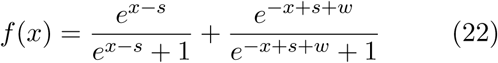

where *s* is the time shift and *w* is the time window of the control measure; both are measured in days. The time dependent control measure function is *λ*(*t*) = *f* (*t*).

Because the rectangular function has a range [1,2], we re-scale the function by subtracting the function by 1 so its range becomes [0, 1].

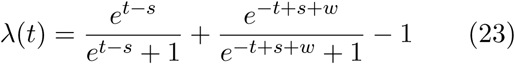

### 4.1 Result from New Jersey

While the SIR model helps simulate the susceptible, infected, and recovering populations, the population that is required to match the actual New Jersey cases is the new cases population. To account for this, as seen in (Figure 13), an array of new cases is created by taking the one day’s population and subtracting the previous day’s population. This creates a graph that illustrates the number of new coronavirus cases each day, rather than the cumulative infected population each day. In this simulation, N is set to 8.882 *×* 10^6^, representing the population of New Jersey. Generally, this simulation’s graph follows a similar shape as the real data in (figure NJCases). The SIR model equations used to model the infected population in New Jersey are altered in this simulation to assist in graphing a more accurate representation of the New Jersey cases. For example, the variable “shift” is used to account for any delays in the effect of control measures. While control measures may have been put in place in New Jersey, such as individuals quarantining themselves, earlier runs of this simulation show that simulating the population where control measures took effect immediately do not produce accurate results. Subsequently, “shift” is used to test different lengths of times for control measures to take effect. In the end, when shift is set to 15, in addition to the adjustment of all other variables, the graph produced shows more similarities with the actual trends in the new cases population. This trial successfully simulates the real conditions and effects of the coronavirus; at the start of the 100 days, the confirmed cases rise at similar rates and reach a maximum of around 4,000 cases. However, after the graphs reaches their peak, the simulated new cases decrease at a much faster rate than the actual new cases. This could mean that *α*, the effect of death, used in the simulation is too large. If *α* decreases, mortality numbers will decrease and the population will decrease less rapidly. However, if *α* is adjusted in this simulation, this will also affect the section of the graph where the population is increasing. Another possible reason for the difference in slope for the decreasing infected population between the real data and the simulation is that *λ*, representing the control measures, is too large. If *λ* decrease, more people will become infected with more control measures; as a result, the infected population will decrease at a slower rate.

**Figure 13:**
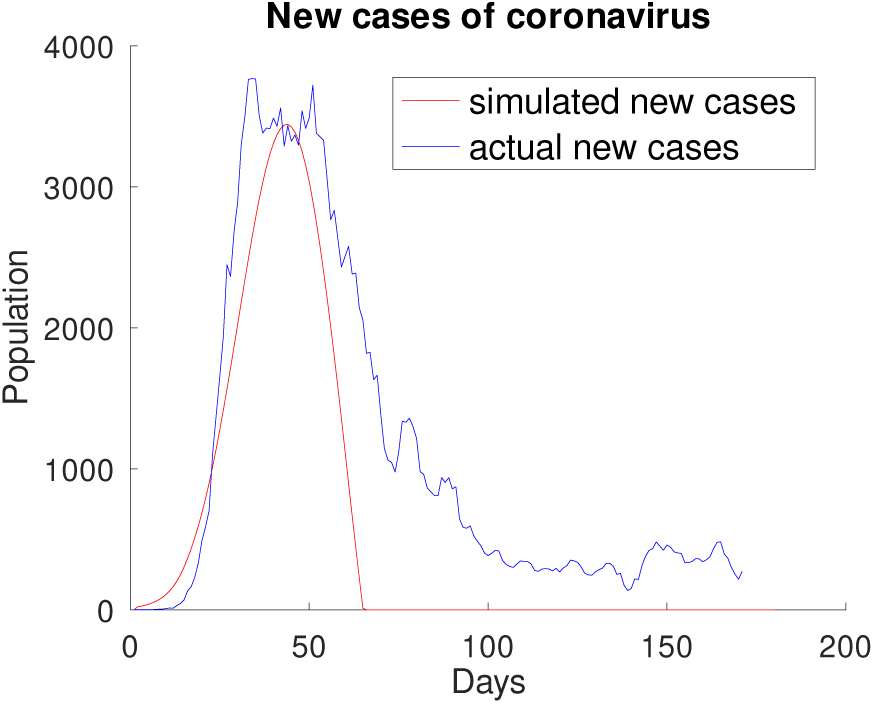
Simulated result of new coronavirus cases in New Jersey using SIR model. Based on the data seen in (Figure 10), variables of the SIR model are adjusted to simulate the rate that the New Jersey population becomes infected over a span of 100 days. To plot this graph, N=8.882 10^6^, *β*=0.28, *γ*=1/14, *λ*=0.026, *α*=0.0001, and shift=15 days.

### 4.2 Result from the U.S

The data in this graph represents 5 day rolling averages of new coronavirus cases each day. This graph shows a rising number of infected individuals starting from around March 27 for about half a month. The graph then reaches a peak, where the new cases continue to fluctuate in growth, but overall show a slight negative slope, meaning the new cases are decreasing. However, around June 27, the new cases begin to grow rapidly once again for about one month. Around July 27, the cases reach a second peak and begin to decrease once again, but this time at a faster rate than the first phase of decrease.

To simulate the new cases population rather than the cumulative infected population that the SIR model helps to graph, a ‘newcase’ array is created, similar to the one created for the New Jersey simulation. In the simulation, (Figure 15), N is set to 3.282 *×* 10^8^, representing the US population. Variables from the SIR model equations are then adjusted to match the actual new cases graph. *β* is set to 0.14, *γ* to 1/200, *λ* to 0.02, and *α* to 0.002. A shift variable is also created for this simulation, similar to the New Jersey simulation. The variable is set to 10, meaning the simulation models new corona virus cases when control measures take effect ten days list. One major difference between this simulation and the New Jersey simulation is the use of two new variables, quarantine window and 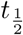 representing how long the control measure is in effect and the time decay constant after which people are more likely to stay in a recovered state, respectively. The addition of these two variables along with the utilization of the rectangular function is what forms the graphs with two peaks. Generally, the simulation results successfully model the real life situation because the number of new cases increases until it reaches a peak, decreases to around 3000 cases until day 100, and then increases again.

**Figure 14:**
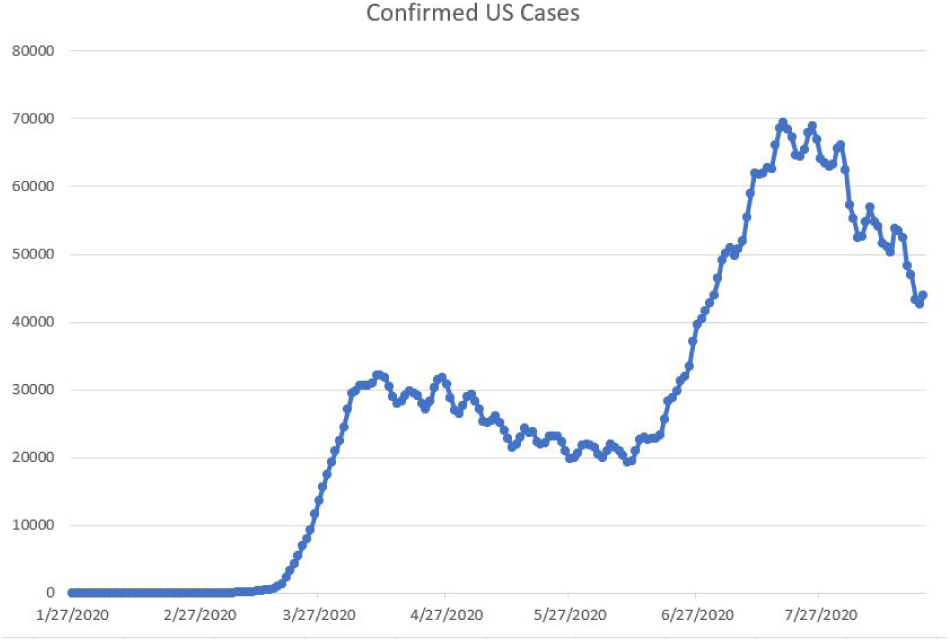
Graph of confirmed coronavirus cases in the United States from 1/27/20 to 8/21/20.

**Figure 15:**
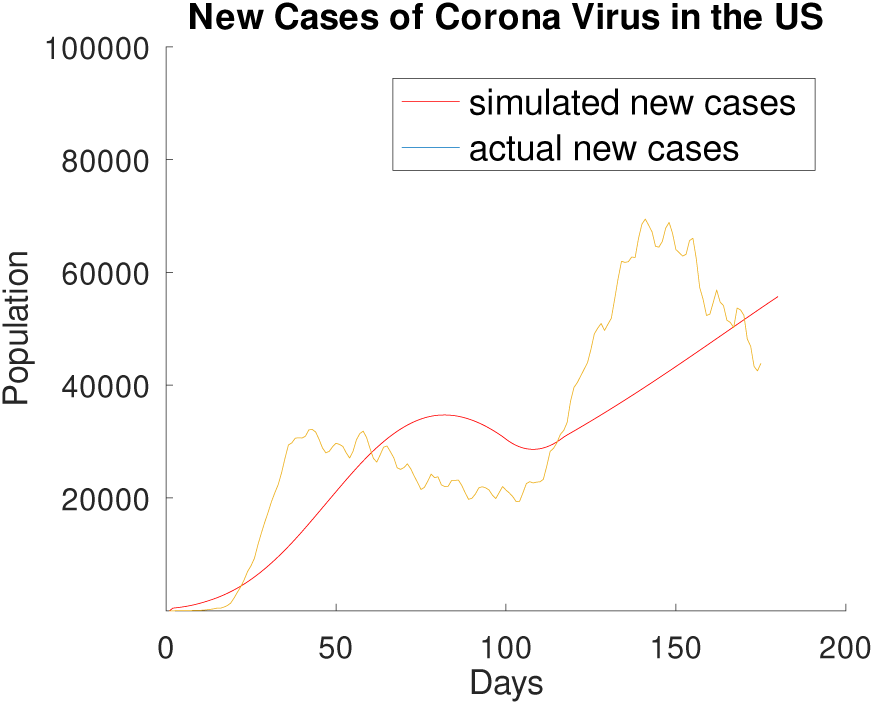
Simulated result of new corona virus cases in the US using the SIR model. As the data of confirmed corona virus cases in the United States produces a graph with two peaks, seen in (Figure 14), a simulation of this span of 7 months is done by adjusting variables from the SIR model equations. To plot this graph, N=3.282 *×* 10^8^, *β*=0.14, *γ*=1/200, *λ*=0.02, *α*=0.002, shift=10 days, quarantine window=95 days, and 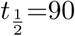 days.

However, as mathematical models have its limitations in accurately simulating real life situations, there are a couple of differences between the simulation and the actual graph of new cases. Although the shapes of the two graphs are similar, the slopes of increasing and decreasing new cases are quite different. For example, the actual new cases increase much more rapidly before the first peak than the simulation. This causes the simulation’s graph to show a peak around 25 days later than the actual graph, showing a delay in the rate that individuals are becoming infected. Moreover, while both graphs show a change from decreasing new cases to increasing new cases a little after 100 days, the actual graph shows a much larger slope. By 150 days, the actual new cases reach approximately 7,000 while the simulation reaches only 5,000 during that time and continues to increase afterwards. By day 180, the actual population of new cases has decreased to about 4,000 cases while the simulation suggests that the number of new cases is still growing.

Several factors can explain the differences between these two graphs. First, limitations of the rectangular function used to simulate the US cases can address the continual growth of new cases after 180 days in Figure 15. While the rectangular function produces a graph with two peaks, it restricts the graph from decreasing after the new cases reach its second peak. Moreover, the value of *λ* used in this simulation may be too large, as the graph generally reaches its peaks much slower than the actual graph of new cases. If *λ* decreases, the effect of control measures will be less noticeable, and more people may become infected, increasing the slope of the rising number of cases before each peak in the graph. Moreover, *β* may be too small in the simulation; if *β* increases, the rate at which individuals become infected will increase, also increasing the slope of rising cases. However, similar to the New Jersey new cases simulation, if both of these variables increases, the shape of the trends in new cases will change and model the actual new cases less similarly.

## 5 Conclusion

The lethal nature of COVID-19 virus has attracted a lot of attention and research work dealing with various aspects of this potent virus. Our work focuses on comparing the new case patterns between the state of New Jersey and the entire US. It’s interesting to understand how the new case patterns between New Jersey and the entire US can be so different.

First, we start by analyzing the behavior of the standard SIR model and examining how the SIR populations depend on the parameters used in the model.

Then, we examine the differences in observed new case patterns between New Jersey and the US. This leads to our conclusion that the difference in new case patterns is mostly caused by the control measure differences between the two regions studied.

To simulate the observed pattern, we introduce time dependent control measure functions into the modified SIR model including additional effects such as secondary infection and an additional mortality population pool along with its associated mortality rate. For the new cases in New Jersey, a step function is used to model the control measure; the control measure taken in New Jersey persists after it takes effect. For the new cases in the US, a rectangular function is used to model the transient nature of the control measure, which is set to last 3 months before it wears off. The difference in the control measure leads to dramatic differences in both observed pattern and simulated patterns as seen in Figure 13 and Figure 15.

As our simulations highlight the importance of quarantine windows, the amount of time that control measures are in effect, it is interesting to note additional work that studies how lifting control measures at specific rates helps to reduce infection rates. Prem and her group [14] simulated lifting control measures such as allowing people to go to work in phases, or a staggered manner; results demonstrated that if the Wuhan population followed these procedures, by mid-2020, the median number of infections would reduce by over 92% [14]. Despite this work that illustrates the benefits of slowly lifting control measures, however, it is still logical to conclude that implementing control measures for a longer period of time would be the safest method of reducing corona virus cases. Further work by Lin and his team demonstrate that social distancing effects are better than traffic control in China, but the combination of both measures would have the most beneficial impact on China’s population regarding corona virus cases [15]. Other work details potential technological methods of control measures; Ferretti explored the feasibility of protecting the population by studying isolation strategies paired with contact tracing [16]. Based on their research results, the team concluded that a possible solution to reduce corona virus cases was to develop an application to allow for instant contact tracing. Through this app, when an individual is diagnosed with COVID-19, their contacts are instantly, automatically, and anonymously notified of their risk and prompted to quarantine themselves. The implementation of such technology could significantly reduce the rate at which individuals become infected with the virus.

These results strongly suggest that the effectiveness of control measures, such as quarantine and wearing masks, should not be overlooked. With persistent control measures, New Jersey new cases decline sharply, while a lack of control measures leads to second wave of growth of new cases in the entire US. As of now, the populace is more aware of these control measure effects and wearing masks has become the standard procedure.

## Data Availability

Data is available from JHU COVID-19 github:
https://github.com/CSSEGISandData/COVID-19

